# Role of contralesional cortico-reticulospinal tract compensation in walking recovery after stroke

**DOI:** 10.64898/2025.12.11.25341794

**Authors:** Jolene Foster, Oluwole O. Awosika, Pierce Boyne

**Author notes:** **Corresponding Author:** Jolene Foster, PT, DPT, NCS, Department of Rehabilitation, Exercise and Nutrition Sciences University of Cincinnati.

## Abstract

**Purpose:** Evidence suggests the contralesional cortico-reticulospinal tract (cCRST) upregulates after stroke, and that this upregulation correlates with worse motor function, suggesting it may be harmful for walking recovery. However, this relationship may be confounded by the extent of ipsilesional corticospinal tract (CST) and CRST damage, which could cause both greater cCRST upregulation and worse walking function. No previous studies have tested whether this confounding relationship exists, nor whether the amount of damage to the ipsilesional motor tracts is related to the amount of cCRST upregulation. We hypothesized that lower ipsilesional motor tract strength would: (1) be associated with greater cCRST compensation; and (2) explain the negative association between cCRST compensation and worse walking function.

**Methods:** Ten individuals with chronic stroke and ten age- and sex-matched controls completed diffusion MRI, from which quantitative anisotropy was derived to evaluate the strength of the ipsilesional and contralesional CRST and CST. Walking capacity was assessed using 6-minute walk distance (6MWD). Linear regressions were applied to examine relationships among ipsilesional corticomotor tract strength (iCRST and iCST combined), cCRST strength, and 6MWD.

**Results:** Compared with controls, participants with stroke had lower ipsilesional and higher contralesional strength for both motor tracts. Lower ipsilesional tract strength was associated with greater cCRST strength z-score (–0.12 SDs [–0.23, –0.02]). The unadjusted association between greater cCRST strength z-score and lower walking capacity (−72 meters [−136, −9]) was no longer present after adjusting for ipsilesional tract strength (−3 meters [−28, 23]).

**Conclusions:** Greater damage to ipsilesional motor tracts (lower strength) was associated with increased cCRST strength. The extent of ipsilesional tract injury fully explained the negative association between cCRST strength and worse walking capacity. These findings suggest that cCRST upregulation is an adaptive compensation mediated by the extent of ipsilesional tract damage, and unlikely to impede walking recovery.

## INTRODUCTION

Despite targeted locomotor interventions, an estimated 80% of patients still experience limitations in walking function after stroke,^1^ including reduced walking capacity (i.e. speed and endurance), which affects their ability to participate in daily activities.^2,3^ A better understanding of how brain pathways reorganize in relation to walking capacity after stroke may lead to improved patient outcomes by guiding interventions such as non-invasive brain stimulation (NIBS), which enhance neural mechanisms involved in recovery. While NIBS has shown promise for improving walking outcomes after stroke, more evidence is needed to support decision-making on which hemisphere and neural tracts to target, and whether to facilitate or inhibit those targets to support improved walking outcomes.^4,5^

The corticospinal tract (CST) is traditionally considered the primary descending motor tract driving functional movement in humans, however, patients with complete damage to the CST have demonstrated the ability to recover independent walking.^6–10^ These findings suggest other non-CST neural tracts can contribute to walking recovery after stroke. Accumulating evidence suggests the cortico-reticulospinal tract (CRST) may play a central role in post-stroke walking recovery. For example, white matter integrity of the ipsilesional CRST (iCRST) appears to be strongly correlated with walking independence and capacity.^11,12^ Unlike the CST, the CRST also projects more bilaterally to the brainstem and spinal cord and is mainly ipsilateral, providing an anatomical basis for the contralesional hemisphere to compensate for ipsilesional tract damage.^13^

Evidence also suggests that the contralesional CRST (cCRST) may be upregulated after stroke,^7,12,14^ but it remains unclear whether this is helpful or harmful. While greater cCRST strength (i.e. upregulation) has been associated with worse motor function,^14^ this association could be confounded by stroke severity rather than reflecting a maladaptive role for cCRST upregulation. In other words, this association could be explained by greater ipsilesional damage causing both greater compensatory cCRST upregulation and worse motor function. This aligns with the bimodal-balance recovery model proposed by Di Pino et al.,^15^ which suggests that the brain’s response to stroke depends on the extent of residual structural integrity in ipsilesional motor pathways. According to this model, individuals with higher structural reserve (i.e. less damage) are more likely to recover through restoration of ipsilesional motor pathways, while those with lower structural reserve (i.e. more damage) may rely more on compensatory mechanisms involving the contralesional hemisphere.^15^ In this context, cCRST upregulation may represent an adaptive, compensatory response in individuals with more severe ipsilesional damage, rather than a maladaptive response.

Some supporting evidence for this can be found in one study that limited the sample to only those with complete ipsilesional motor tract damage, partly controlling for this confounding. In this study, greater cCRST upregulation was indeed associated with a higher motor function and greater likelihood of achieving independent walking.^7^ However, no previous studies have tested whether this confounding relationship exists, or whether the extent of motor tract damage is associated with the amount of cCRST upregulation. Understanding this relationship is essential for determining whether cCRST upregulation is a compensatory response and could provide insights to inform future interventions.

This study aimed to assess whether cCRST upregulation is a compensatory response by testing: (1) whether cCRST upregulation magnitude relates to ipsilesional motor tract strength; and (2) whether the relationship between cCRST upregulation and walking function is confounded by ipsilesional motor tract strength. We hypothesized that lower ipsilesional motor tract strength would: (1) be associated with greater cCRST compensation; and (2) explain the negative association between cCRST compensation and worse walking function.

## METHODS

### Study Design

The study providing data for this analysis was approved by institutional review boards, preregistered on ClinicalTrials.gov (NCT02858349) and performed in a cardiovascular stress laboratory, magnetic resonance imaging (MRI) research center, and rehabilitation research laboratory from July 2016 to December 2017. Some of the methods and baseline data from this study have been previously described in manuscripts addressing different aims from the current report.^16–18^

### Participants

Ten participants with chronic stroke and 10 age- and sex- matched controls were recruited from the community and provided written informed consent. Inclusion criteria for both groups were: age 30–90 years old, MRI compatible, able to communicate with investigators and answer consent comprehension questions correctly, able to perform mental imagery,^19^ no recent history of drug/alcohol abuse or significant mental illness, and not pregnant. Additional inclusion criteria specifically for participants with stroke were: unilateral stroke >6 months prior to enrollment in middle cerebral artery territory; walking speed <1.0 m/s on the 10 meter(m) walk test,^20^ able to walk 10m with an assistive device as needed and no physical assistance by another person; no evidence of significant arrhythmia or myocardial ischemia on treadmill electrocardiogram (ECG) stress test, no significant baseline ECG abnormalities that would make an exercise ECG uninterpretable;^21^ no recent cardiopulmonary hospitalization; no significant ataxia or neglect (NIHSS item score > 1);^22^ no severe lower extremity (LE) hypertonia (Ashworth > 2);^23^ no major post-stroke depression (PHQ-9 ≥ 10)^24^ in the absence of management by a health care provider;^25^ not participating in physical therapy or another interventional research study; no recent paretic LE botulinum toxin injection; and no other progressive neurologic disorder or other major conditions that would limit capacity for improvement. Control participants were a demographic match for a participant with stroke (sex, age difference ≤ 5 years) without any current neurologic, orthopedic, or medical condition affecting walking function.

### Walking Capacity Assessment

Walking capacity was measured by the 6-minute walk distance (6MWD). During the test, the participant is instructed to walk as far as possible in 6 minutes using a standardized course.^26^ The distance walked provides a reliable and valid measure of walking capacity and is associated with community ambulation after stroke.^27,28^ A blinded rater assessed the test.

### MRI Data Acquisition

A 3.0T Philips Ingenia MRI system was used. T1-weighted brain images were acquired at 1mm isotropic resolution with the following parameters: TR, 8.1ms; TE, 3.7ms; flip angle, 8^0^; SENSE factor 2. Diffusion weighted brain images were acquired at 2mm isotropic resolution with the following parameters: TR, 7.1s; TE, 92ms; flip angle, 90^0^; 61 directions at b=1000 s/mm^2^; 7 b=0 volumes; SENSE factor 3.

### MRI Data Preprocessing

For the T1 images, FSL software^29^ was used for bias field correction, tissue type segmentation and non-linear registration to the MNI152 template, using a lesion mask to improve registration for participants with stroke by temporarily filling the lesion with MNI template voxels.^30^ Diffusion MRI preprocessing included eddy current correction, motion correction, and outlier volume replacement using FSL’s “eddy” tool.^29,31,32^ Alignment to the T1 image with boundary-based registration was performed using the T1 white matter segmentation.^29,33^ Diffusion data were reconstructed with generalized Q sampling imaging using DSI Studio (https://dsi-studio.labsolver.org/) with a diffusion length ratio of 1.7, to calculate quantitative anisotropy (QA) in each brain voxel.^34,35^

Diffusion anisotropy metrics such as fractional anisotropy (FA) and QA are commonly used as markers of tract strength to assess residual microstructural integrity and neural reorganization processes after stroke.^36–38^ Lower anisotropy is associated with greater white matter damage and motor impairment, whereas higher anisotropy reflects better microstructural integrity, organization, and axonal remodeling.^37–40^ While FA is the most widely used anisotropy metric, it has limitations. For example, FA is sensitive to extracellular water (i.e. edema) and multiple fiber directions within a voxel, both of which make FA values underestimate axonal projection strength.^36^ In contrast, QA is less susceptible to partial volume effects and offers improved resolution of multiple fiber directions, making it a more specific indicator of axonal projection strength.^36^ In this study, we use QA of ipsilesional motor tracts as a measure of ipsilesional motor tract damage and QA of the contralesional motor tracts to quantify neuroplasticity (i.e. contralesional compensation).

### Voxel-based Normalization

To reduce the impact of scanner variability, QA data were normalized (nQA) by dividing the QA value in each voxel by the mean QA within the ventricular cerebrospinal fluid (CSF) from the same scan.^41^ CSF was selected as the normalization reference because QA values in CSF are not influenced by disease pathology.^41^

nQA values were projected onto a standard space map of the core white matter “skeleton” using a method analogous to FSL’s Tract-Based Spatial Statistics (TBSS).^42^ This approach helps correct residual misalignments after standard space registration, which is important after stroke because of tissue changes that complicate registration.^43,44^ For each voxel in the white matter skeleton, the highest nearby nQA value for the participant was projected onto the skeleton, to better align the center of each white matter bundle to the standard space template.^42^

### Tract Strength Measurement

For each participant, mean nQA values were calculated within each tract (iCRST, cCRST, iCST, cCST). The contribution of each voxel to the mean nQA value was weighted by the number of streamlines passing through that voxel in the normative template for that tract.^45^ This calculation specifically used voxels in the internal capsule region (z = −5 to 12 mm) of each tract, because that region is less prone to diffusion MRI distortions^46^ and anisotropy measures show the strongest association with motor outcomes (i.e. greatest concurrent validity) when extracted from that region.^40^

The nQA measurement for each tract was then adjusted for the global mean cerebral white matter nQA for that participant, to more specifically quantify the strength of that tract, independent from global white matter strength. To do this, linear regression models were developed using control participant data to quantify the relationship between global nQA (independent variable) and tract-specific nQA (dependent variable), averaged across the left and right tract. These models had regression slopes (β) of 1.186 and 2.700 and r^2^ values of 0.735 and 0.883 for the CRST and CST, respectively. The regression slopes from these models and the mean global nQA across the control participants (2.662) were then used to adjust the tract nQA values for each participant, using the following formulas for both the ipsilesional and the contralesional tracts (the ipsilesional tract for controls was the same as their matched counterpart with stroke for all formulas):

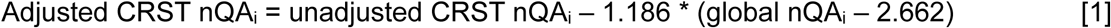

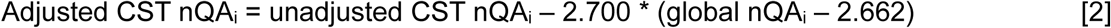

In addition to calculating separate strength measures for each tract, we also calculated overall corticomotor strength for each hemisphere by averaging the CRST and CST values. The contribution of each tract to this average was weighted by the mean normative volume of that tract, to account for differences in tract size.^45^ Finally, to improve interpretability, tract nQA values were then converted to z scores by subtracting the control mean and dividing by the control standard deviation, using the following formulas:

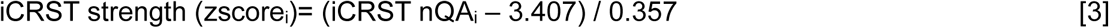

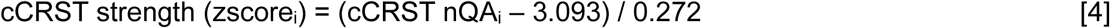

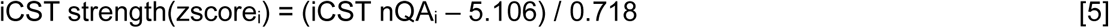

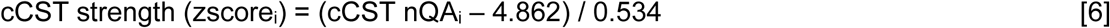

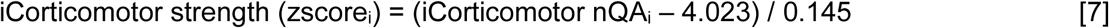

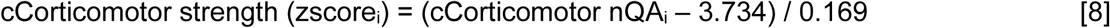

### Data Analysis

Participant characteristics and tract strength values were compared between participants with stroke and controls using independent t-tests. To address the hypotheses, linear regressions were applied to assess relationships between iCorticomotor strength, cCRST strength and 6MWD. To assess whether contralesional cCRST upregulation is related to ipsilesional motor tract strength, the following regression was used:

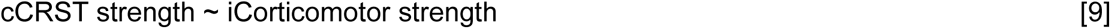

To assess whether the negative association between cCRST projection strength and 6MWD is confounded by ipsilesional motor tract strength, the following regressions were used:

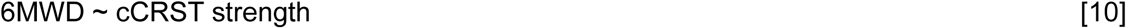

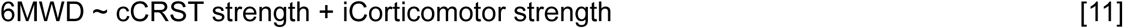

Confounding by the ipsilesional motor tract strength was determined to be present if adding iCorticomotor strength to the model shrunk the regression coefficient for cCRST strength by more than 10% (from model 10 to 11).^47^ All analyses were conducted using R version 4.4.1 (2025.9.0.387).^48^

### Sample Size Calculation

The target sample size of 10 participants with stroke and 10 controls was calculated for different aims than those reported here.^17^ That sample size provides 80% estimated power to detect: 1) a within-group Cohen’s d effect size as small as 1.00; 2) a between-group effect size as small as 1.32; and 3) a correlation as low as 0.58. These calculations were based on a two-sided significance level of 0.05 and were performed with the R package ‘pwr’.^17^

## RESULTS

The target sample size (10 participants with stroke and 10 controls) was reached, and there were no missing data. Compared with controls, participants with stroke had significantly lower comfortable gait speeds and 6-minute walk distances, but similar age, and body mass index (Table 1).

**Table 1.** Participant characteristics.

|  | Stroke<br>(N=10) | Control<br>(N=10) | Stroke vs. control<br>p-value |
| --- | --- | --- | --- |
| Age (years) | 59.8 ± 6.78 | 58.9 ± 7.79 | 0.79 |
| Gender, N (%) |  |  |  |
| Female | 4 (40.0%) | 4 (40.0%) |  |
| Male | 6 (60.0%) | 6 (60.0%) |  |
| Body Mass Index (kg/m<sup>2</sup>) | 30.2 ± 4.24 | 30.8 ± 4.25 | 0.76 |
| Comfortable gait speed (m/s) | 0.41 ± 0.33 | 1.36 ± 0.16 | <0.0001 |
| Six-minute Walk distance (m) | 156 ± 147 | 537 ± 77.1 | <0.0001 |
| Years since stroke | 2.4 ± 1.7 | NA |  |
| Type of stroke, N (%) |  |  |  |
| Hemorrhagic | 1 (10.0%) | NA |  |
| Ischemic | 9 (90.0%) | NA |  |
| Lesion side, N (%) |  |  |  |
| Left | 5 (50.0%) | NA |  |
| Right | 5 (50.0%) | NA |  |
| Lesion volume, mL | 122 ± 105 | NA |  |
| Assistive device, N (%) |  |  |  |
| Hemiwalker | 1 (10.0%) | 0 (0%) |  |
| Wide based quad cane | 4 (40.0%) | 0 (0%) |  |
| Narrow based quad cane | 2 (20.0%) | 0 (0%) |  |
| None | 3 (30.0%) | 10 (100%) |  |
| Orthotics, N (%) |  |  |  |
| AFO | 8 (80.0%) | 0 (0%) |  |
| None | 2 (20.0%) | 10 (100%) |  |

Values are mean ± SD or N (%). P-values are from independent T-tests. NA, not applicable.

Significant between-group differences in motor tract strength were found for all four motor tracts assessed. When compared to controls, participants with stroke demonstrated decreased motor tract strength of the iCRST and iCST and increased strength of the cCRST and cCST (Table 2 and Figure 1).

**Figure 1.**
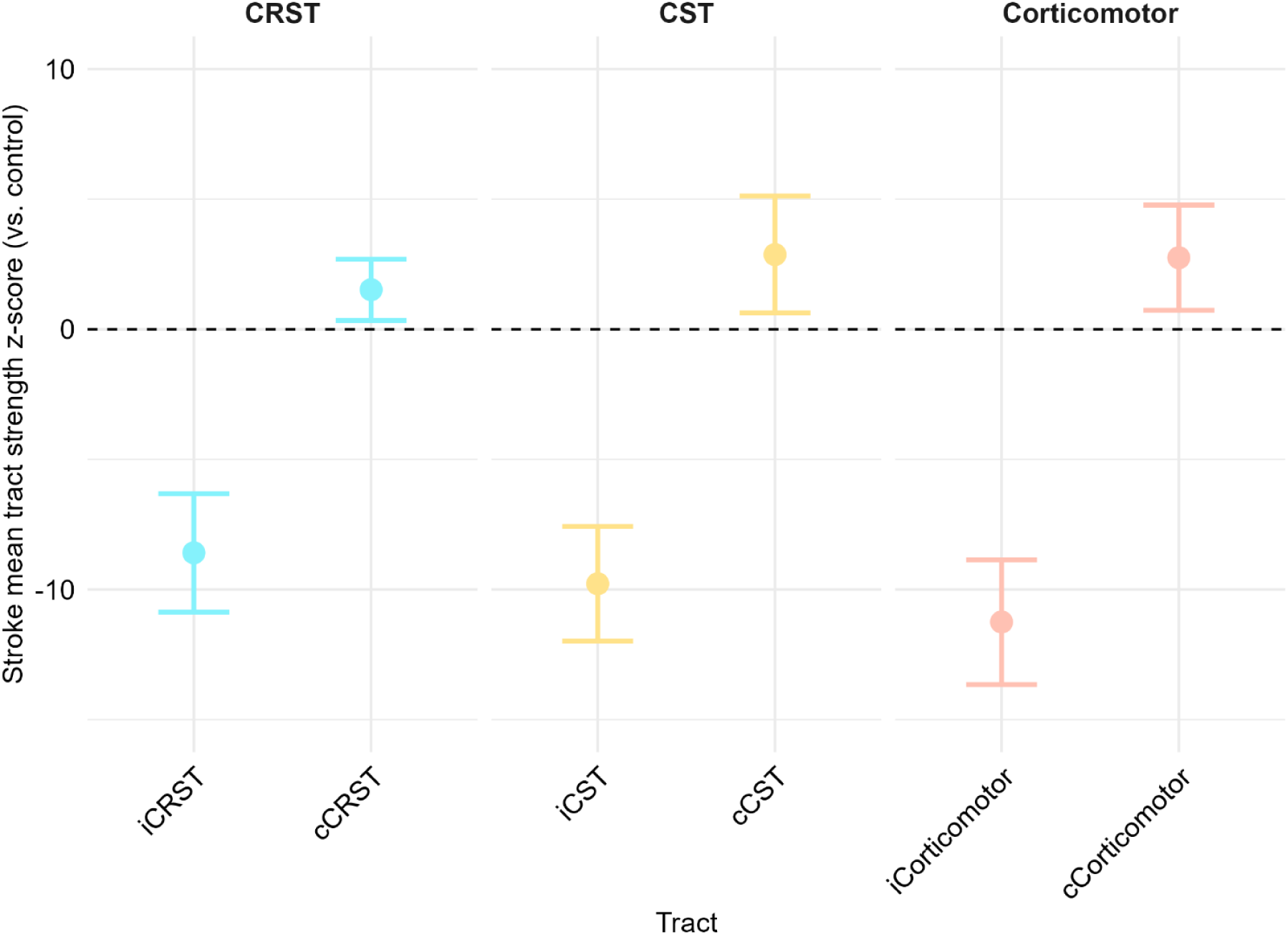
Stroke motor tract strength z-scores relative to controls. CRST, cortico-reticulospinal tract; iCRST, ipsilesional cortico-reticulospinal tract; cCRST, contralesional cortico-reticulospinal tract; CST, corticospinal tract; iCST, ipsilesional corticospinal tract; cCST, contralesional corticospinal tract; iCorticomotor, combined iCRST and iCST; cCorticomotor, combined cCRST and cCST; Stroke z-scores were calculated based on the mean and standard deviation in the control group and represent the number of standard deviations above or below the control mean value for that tract.

**Table 2.** Stroke vs. control motor tract strength.

| Tract | Stroke (N=10),<br>nQA | Control (N=10),<br>nQA | Stroke – control,<br>nQA | Stroke (N=10),<br>z-score |
| --- | --- | --- | --- | --- |
| iCRST | 2.20 ± 0.45 | 3.41 ± 0.14 | -1.21 [-1.52, -0.90] | -8.59 [-10.87, -6.32] |
| cCRST | 3.43 ± 0.37 | 3.09 ± 0.22 | 0.34 [0.05, 0.62] | 1.52 [0.34, 2.69] |
| iCST | 2.73 ± 0.75 | 5.11 ± 0.24 | -2.37 [-2.90, -1.85] | -9.78 [-11.98, -7.58] |
| cCST | 5.55 ± 0.75 | 4.86 ± 0.24 | 0.69 [0.16, 1.21] | 2.87 [0.63, 5.12] |
| iCorticomotor | 2.39 ± 0.49 | 4.02 ± 0.14 | -1.63 [-1.97, -1.29] | -11.25 [-13.65, -8.86] |
| cCorticomotor | 4.20 ± 0.48 | 3.73 ± 0.17 | 0.46 [0.13, 0.80] | 2.75 [0.73, 4.77] |

Normalized quantitative anisotropy (nQA) values for stroke and control groups are mean ± standard deviation QA values as a multiple of the mean QA in the ventricular cerebrospinal fluid. Stroke - control differences are model estimates [95% confidence interval]. Stroke z-scores were calculated based on the mean and standard deviation in the control group and represent the number of standard deviations above or below the control mean value for that tract. CRST, cortico-reticulospinal tract; iCRST, ipsilesional cortico-reticulospinal tract; cCRST, contralesional cortico-reticulospinal tract; CST, corticospinal tract; iCST, ipsilesional corticospinal tract; cCST, contralesional corticospinal tract; iCorticomotor, combined iCRST and iCST; cCorticomotor, combined cCRST and cCST.

When testing whether cCRST upregulation magnitude relates to ipsilesional motor tract strength, linear regression showed a significant negative association between ipsilesional motor tract strength and cCRST strength (−0.12 SDs [−0.23, −0.02]). When testing whether the relationship between cCRST upregulation and walking function is confounded by ipsilesional motor tract strength, the unadjusted linear regression showed a significant negative association between the 6MWD and cCRST strength. This negative association was no longer present when adjusting for ipsilesional motor tract strength (Table 3 and Figure 2).

**Figure 2.**
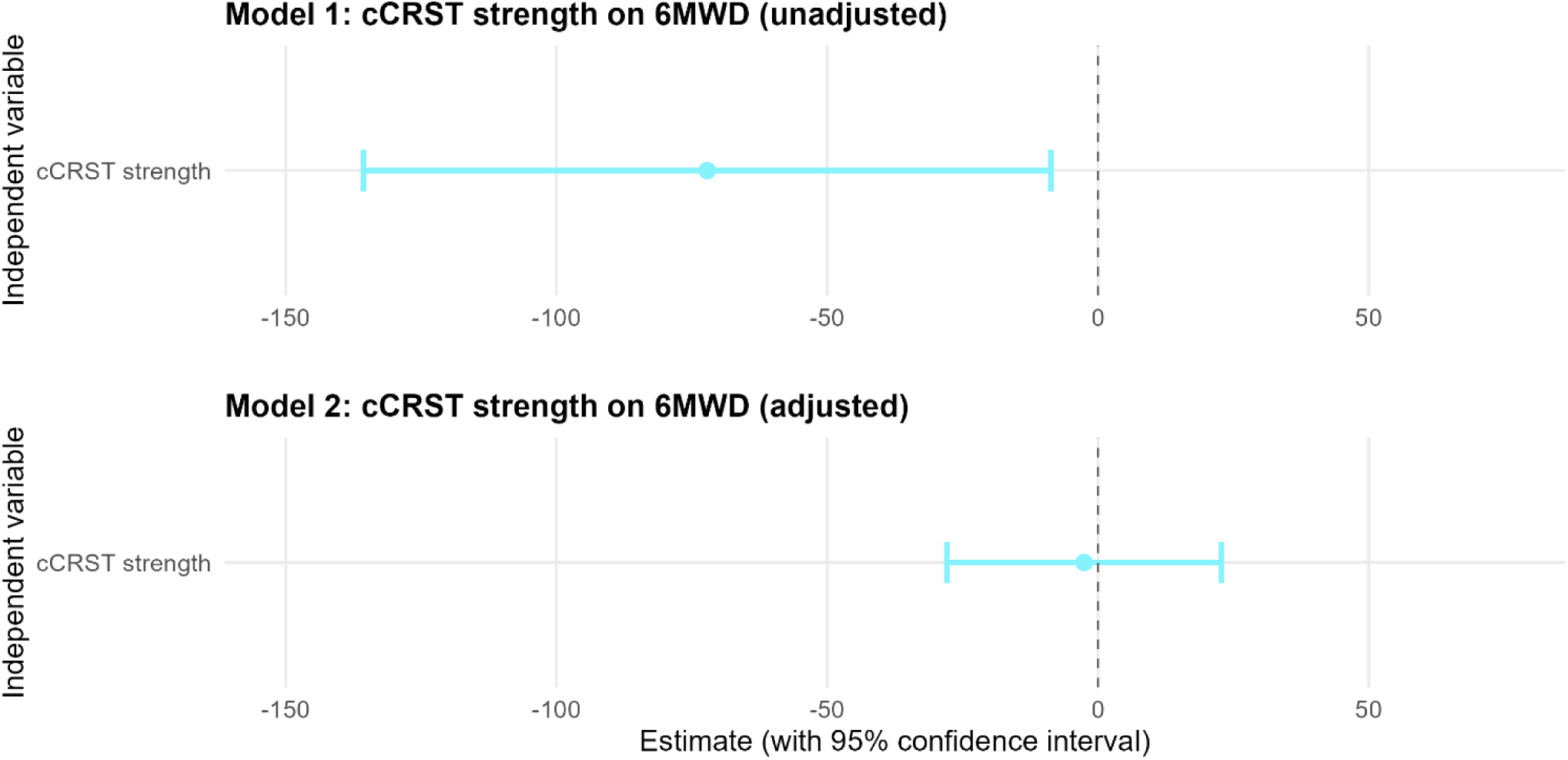
Linear regression of cCRST on 6MWD. cCRST, contralesional cortico-reticulospinal tract; 6MWD, six-minute walk distance

**Table 3.** Linear regression of cCRST strength on 6MWD.

| **Model 1: Unadjusted for ipsilesional damage** |  |  |  |  |
| --- | --- | --- | --- | --- |
| Variable | Estimate | Std. Error | t value | Pr(>\|t\|) |
| (Intercept) | 401.38 | 50.73 | 7.91 | 0.00 |
| cCRST strength | -72.17 | 30.21 | -2.39 | 0.03 |
| Residual standard error: 202.4 on 18 degrees of freedom, multiple R-squared: 0.2407 (adjusted R-squared: 0.1985), F-statistic: 5.706 on 1 and 18 degrees of freedom, p-value: 0.02807 |  |  |  |  |
| **Model 2: Adjusted for ipsilesional damage** |  |  |  |  |
| Variable | Estimate | Std. Error | t value | Pr(>\|t\|) |
| (Intercept) | 541.44 | 21.23 | 25.50 | 0.00 |
| cCRST strength | -2.56 | 12.01 | -0.21 | 0.83 |
| iCorticomotor strength | 34.26 | 2.95 | 11.61 | 0.00 |
| Residual standard error: 69.72 on 17 degrees of freedom, multiple R-squared: 0.9149 (adjusted R-squared: 0.9049), F-statistic: 91.43 on 2 and 17 degrees of freedom, p-value: 7.994e-10 |  |  |  |  |
| cCRST, contralesional cortico-reticulospinal tract; 6MWD, six-minute walk distance; Intercept, 6-minute walk distance when variables = 0; iCorticomotor strength; the sum of strength of the ipsilesional corticoreticulospinal tract and ipsilesional corticospinal tract. |  |  |  |  |

## DISCUSSION

This study sought to clarify whether cCRST upregulation represents a compensatory response to stroke. Compared with age and sex-matched controls, participants with stroke had significantly lower strength of the ipsilesional motor tracts (iCorticomotor, iCRST, and iCST), and significantly higher strength of the analogous tracts in the contralesional hemisphere. The lower strength of ipsilesional tracts presumably reflects damage from the stroke, while the higher strength of contralesional tracts indicates upregulation of those pathways.

When assessing whether cCRST upregulation magnitude relates to ipsilesional motor tract strength, we found that lower ipsilesional strength was significantly associated with greater cCRST strength. This finding is consistent with the possibility that cCRST upregulation is a compensation for ipsilesional motor tract damage. The predominant ipsilateral projections from the cCRST to the paretic side of the body provide an anatomical substrate for this potential compensatory role.^13,45^ Our findings may help explain the cCRST upregulation observed in prior studies^7,12,14^ and extend prior knowledge by demonstrating a scaled relationship between the extent of ipsilesional damage and cCRST upregulation.

We also found evidence that the association between cCRST upregulation and walking function is confounded by the residual strength of the ipsilesional motor pathways. Similar to Srivastava et al.^12^, our unadjusted model showed that greater cCRST strength was associated with worse walking capacity (lower 6MWD). However, we found that this negative association was no longer present when adjusting for ipsilesional motor tract strength (see Table 3 and Figure 2), which is the hallmark sign of confounding. This finding directly addresses the ongoing debate over whether cCRST upregulation is adaptive or maladaptive for walking recovery, suggesting that it may not be maladaptive as once thought. Our finding is also consistent with a prior study which found greater estimated cCRST volume among ambulatory vs. non-ambulatory participants.^7^ Considering our current results, this positive association between cCRST upregulation and walking independence in this prior study might have been observed because only individuals with complete damage of the iCST were included. This would partially control for ipsilesional motor tract strength (albeit not in the same way as the current study), allowing more accurate assessment of the effect of cCRST upregulation on walking independence. Taken together with the current results, these findings suggest that cCRST upregulation is not broadly harmful to walking function. Instead, cCRST upregulation may reflect an attempt by the brain to maintain function when the ipsilesional motor system is compromised.

These findings also align with the bi-model balance recovery model proposed by Pino et al.^15^ According to this model, the functional role of the contralesional hemisphere, such as cCRST compensation, depends on ipsilesional motor tract strength.^15^ Our data support this framework by showing that cCRST upregulation scales with the extent of ipsilesional damage, and that cCRST upregulation was not independently associated with poorer walking outcomes after accounting for the extent of damage to the ipsilesional motor tracts. If this model holds with further testing, it suggests the need for individualized neurorehabilitation strategies tailored to each patient and may support targeting contralesional motor pathways in patients with low ipsilesional motor tract strength.

### Limitations

One limitation of this study is limited generalizability due to the small sample size. Additionally, the low mean ipsilesional tract strength and walking capacity in the study sample may limit the generalizability of the findings to individuals with milder stroke. Finally, the cross-sectional design precludes causal inference. Thus, larger longitudinal studies are needed to confirm our findings and further assess the impact of cCRST compensation on walking outcomes in patients after stroke.

### Conclusion

This study demonstrates that greater ipsilesional motor tract damage is associated with increased cCRST strength. We also found that the negative association between cCRST strength and worse walking capacity was explained by the extent of residual ipsilesional motor tract strength. Therefore, our findings suggest cCRST upregulation is not inherently harmful to walking recovery after stroke and may be an adaptive neuroplastic response to maximize function when ipsilesional motor pathways are more damaged.

## Funding

This work was supported by the National Institutes of Health [grant numbers KL2TR001426, UL1TR001425, R01HD093694]; and the American Heart Association [grant number 17MCPRP33670446].

## Data Availability

The data that support the findings of this study are available from the corresponding author upon reasonable request.

## Ethics and consent statement

Written informed consent was obtained from all participants before the experiment, and the study protocol was approved by the University of Cincinnati IRB # 2016-1916.

## Preregistration of Studies

This study was prospectively registered on ClinicalTrials.gov (Identifier: NCT02858349) prior to participant enrollment.

## Author Contributions

JF: Methodology, Software, Formal analysis, Visualization, Writing – original draft, Writing-review & editing. OA: Methodology and Writing-review & editing. PB: Conceptualization, Data curation, Funding acquisition, Investigation, Methodology, Project administration, Resources, Software, Validation, Visualization, Writing – review & editing, Supervision. All authors contributed to the article and approved the submitted version.

## Conflicts of Interest

The authors declare no conflicts of interest.

## Notes

### Competing Interest Statement

The authors have declared no competing interest.

### Clinical Trial

NCT02858349

### Author Declarations

Ethics committee/IRB of the University of Cincinnati gave ethical approval for this work under IRB number 2016-1916.

